# Dynamics of IgG seroconversion and pathophysiology of COVID-19 infections

**DOI:** 10.1101/2020.06.07.20124636

**Authors:** Henry M Staines, Daniela E Kirwan, David J Clark, Emily R Adams, Yolanda Augustin, Rachel L Byrne, Michael Cocozza, Ana I Cubas-Atienzar, Luis E. Cuevas, Martina Cusinato, Benedict M O Davies, Mark Davis, Paul Davis, Annelyse Duvoix, Nicholas M Eckersley, Daniel Forton, Alice J Fraser, Gala Garrod, Linda Hadcocks, Qinxue Hu, Michael Johnson, Grant A Kay, Kesja Klekotko, Zawditu Lewis, Derek C Macallan, Josephine Mensah-Kane, Stefanie Menzies, Irene Monahan, Catherine M Moore, Gerhard Nebe-von-Caron, Sophie I Owen, Chris Sainter, Amadou A Sall, James Schouten, Chris Williams, John Wilkins, Kevin Woolston, Joseph R A Fitchett, Sanjeev Krishna, Tim Planche

**Affiliations:** Centre for Diagnostics and Antimicrobial Resistance, Institute for Infection & Immunity, St George’s University of London, London, UK; Tropical Disease Biology, Centre for Drugs and Diagnostics, Liverpool School of Tropical Medicine, Liverpool, UK; Mologic COVID-19 Diagnostics Development Team; St George’s University Hospitals NHS Foundation Trust, UK; Institut Pasteur de Dakar, Dakar, Senegal; Institut für Tropenmedizin, Universitätsklinikum Tübingen, Tübingen, Germany; Centre de Recherches Médicales de Lambaréné, Gabon

## Abstract

We report dynamics of seroconversion to SARS-CoV-2 infections detected by IgG ELISA in 177 individuals diagnosed by RT-PCR. Longitudinal analysis identifies 2-8.5% of individuals who do not seroconvert even weeks after infection. They are younger than seroconverters who have increased co-morbidity and higher inflammatory markers such as C-Reactive Protein. Higher antibody responses are associated with non-white ethnicity. Antibody responses do not decline during follow up almost to 2 months. Serological assays increase understanding of disease severity. Their application in regular surveillance will clarify the duration and protective nature of humoral responses to SARS-CoV-2.

## Introduction

SARS-CoV-2 is a betacoronavirus that causes Coronavirus Disease-19 (COVID-19), an acute severe respiratory infection with systemic involvement and an estimated case fatality rate of 1%^1^. COVID-19 was first documented in Wuhan (Hubei province) China^2^ at the end of 2019 and rapidly transformed from a localised outbreak to a global pandemic. Countries have tried to manage the pandemic by implementing different strategic interventions with variable success^3^. All agree that diagnostic tests, and the subsequent interventions they unlock, are central to controlling SARS-CoV2 transmission. COVID-19 has introduced formidable disruption in low income countries where diagnostic tests remain unaffordable.

National strategies for containment are severely limited by availability of clinically validated diagnostic tests. The urgent scale up of global nucleic acid amplification test (RT-PCR) capabilities exposed further vulnerabilities in supply chains including shortages of swabs and reagents for extraction and amplification of RNA. RT-PCR became available soon after SARS-CoV-2 was sequenced because such tests do not need production of viral proteins^4^ or assay development that relies on material from COVID-19 patients and those who have never been infected.

Serological assays for viral infections can contribute to all aspects of their containment including vaccine development, diagnostic deployment, and prescription of novel therapeutics. They may also clarify pathophysiological aspects of COVID-19. We have used an enzyme-linked immunosorbent assay (ELISA, Mologic Ltd, Bedford, UK) and rapid diagnostic tests (RDT) for COVID-19 developed for affordability and accuracy so that they can be accessible and can be manufactured in low- and middle-income countries (LMICs). We describe the application of this ELISA to serum samples obtained from a cohort of individuals with confirmed SARS-CoV-2 in London, UK, to identify demographic and clinical variables that may influence dynamics of antibody responses.

## Results

One hundred and seventy-seven patients provided 645 distinct excess diagnostic material (EDM) samples. Their baseline demographic and clinical characteristics are presented in Table 1. Patients were from diverse ethnic backgrounds (34% white, 35% non-white; Supplementary Table 2) with a median (IQR) age of 64 (52-77) years, 57% were male, and most (73%) had one or more co-morbidities. Nineteen percent (34/177) were defined as asymptomatic patients and did not report respiratory symptoms on admission. They were diagnosed with COVID-19 when investigated for other medical conditions. Among the 143 symptomatic patients, the time from symptoms initiation to testing was a median (IQR) of 6 (3-9) days. Ninety four percent (166/177) were hospitalized, the remainder were 7 staff and 4 outpatients; 25% (44/177) died (median (IQR) time to death was 19.1 (14.8-24.8 days)), 61% (108/177) were discharged (median (IQR) length of stay 19.3 (10.6-31.1 days)), and 8% (14/177) were in hospital when the study ended. 36% (63/177) patients needed intensive care.

**Table 1.** Demographic and clinical characteristics of patients.

|  | **Total N = 177 (%)** |
| --- | --- |
| Age, Median (IQR) | 64 (52 - 77) |
| No. of males (%) | 100 (57) |
| BMI, Median (IQR)* (N = 164) | 25.4 (21.9 - 30.5) |
| **Ethnicity** |  |
| White | 60 (33.9) |
| Non-white | 61 (34.5) |
| Other/not known | 56 (31.6) |
| **Comorbidities** |  |
| None | 47 (26.6) |
| 1 co-morbidity | 52 (29.4) |
| 2 co-morbidities | 50 (28.3) |
| 3 or more co-morbidities | 28 (15.8) |
| **Symptoms and diagnostic site** |  |
| Symptomatic | 143 (80.8) |
| Median time from symptoms to RT-PCR swab, days | 6 (3 - 9) |
| Swab taken at |  |
| Emergency department | 90 |
| Outpatients | 12 |
| Wards | 54 |
| Intensive care unit | 14 |
| Occupational health | 7 |
| **Treatment location** |  |
| Occupational Health | 7 (4) |
| Outpatient | 4 (2.3) |
| Hospitalized (%) | 166 (93.8) |
| Admitted to ITU (%) | 63 (38) |
| Discharged (%) | 108 (65.1) |
| Length of stay, days median (IQR) | 19.3 (10.6 - 31) |
| Deceased (%) | 44 (26.5) |
| Length of stay, median (IQR) | 19.1 (14.8 - 24.8) |
| Deceased and/or ITU admission (%) | 80 (48.2) |
| Still in hospital (%) | 16 (9.6) |

\*Height unavailable for 13 patients

Figure 1a summarises normalised optical density (NOD) values from the ELISA, proportional to specific IgG antibodies of patients by time after diagnosis. 8.5% (95%CI 5.2% - 13.5%; 15/177) did not seroconvert during the entire follow up period, 7.3% (95%CI 4.3% - 12.1%; 13/177 seroconverted after enrolment and 84% (95%CI 78% - 89%; 149/177) had already seroconverted at the time of the first serological test. Twenty six percent (4/15) of the non-seroconverters were followed beyond 20 days, suggesting that 2 - 8.5% of patients may not develop detectable IgG antibody responses to SARS-CoV-2 for weeks following infection.

**Fig. 1.**
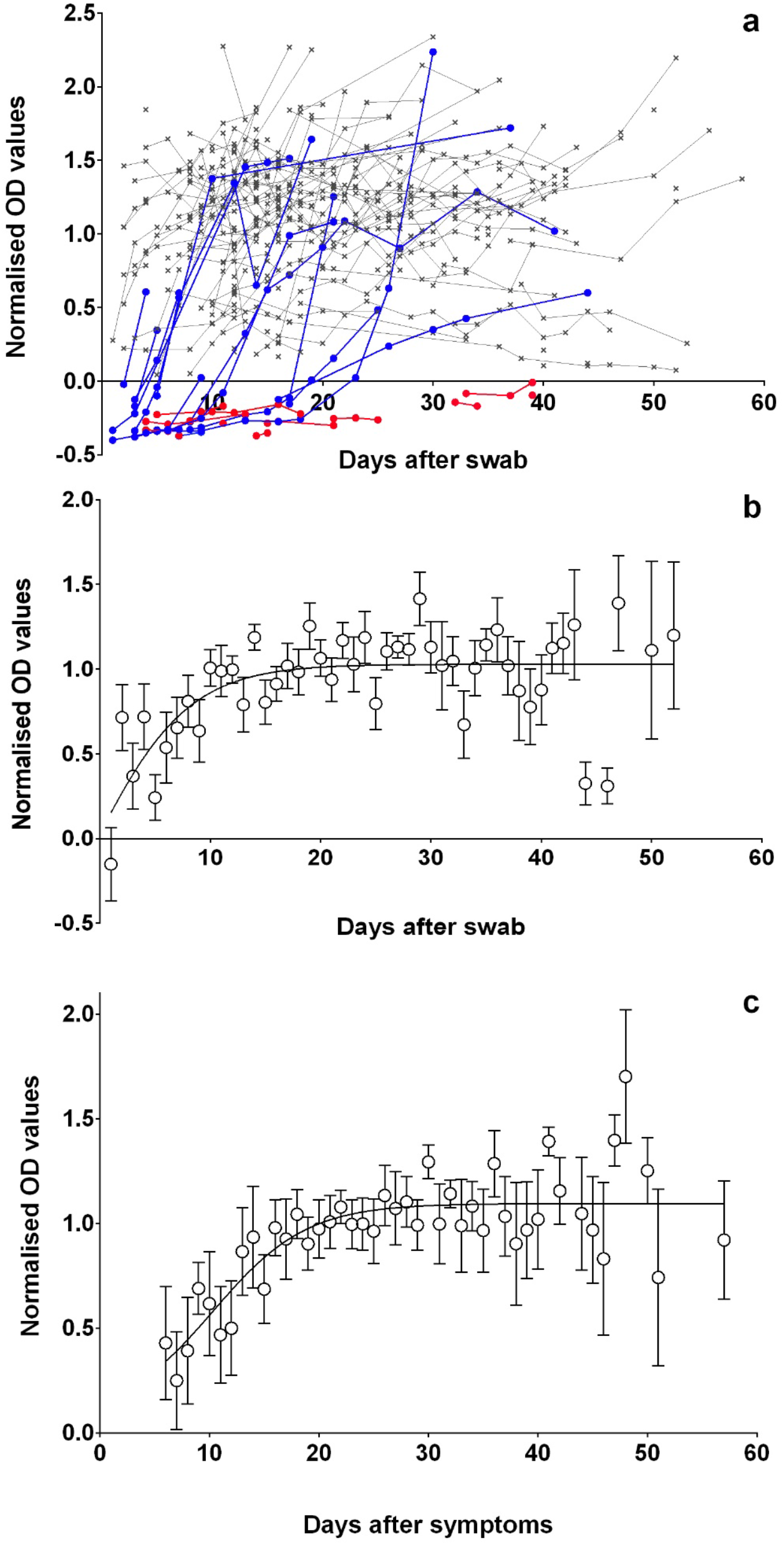
Antibody dynamics, using normalised OD (NOD) values. **a**, Sequential NOD values by days after first RT-PCR positive swab for seroconverted (grey crosses), seroconverting (blue circles) and non-seroconverting patients (red circles). **b**, Mean (± SEM) NOD values (≥ 3 per time point; n = 48) by days after first PCR positive swab for those who seroconverted. A 4-parameter sigmoidal unconstrained model was fitted (r^2^ = 0.45). **c**, Mean (± SEM) NOD values (≥ 3 per time point; n = 45) by days after first symptoms for those who seroconverted. A 4-parameter sigmoidal unconstrained model was fitted (r^2^ = 0.63).

Figures 1b and 1c present NOD IgG values after a positive RT-PCR test and after symptom onset respectively. The difference in the estimated time for NOD values to plateau from RT-PCR (∼12 daysm Fig. 1b) and after onset of symptoms (∼19 daysm Fig. 1c) is ∼7 days and is consistent with the median time of 6 days between symptoms and RT-PCR testing. Following seroconversion, mean NOD values remained stable over the time of study (up to ∼2 months after onset of symptoms).

We next assessed whether if the rate of seroconversion was higher in participants ≤ 70 y and > 70 y, by sex, or the presence of respiratory symptoms. None of these variables discernably influenced seroconversion rates (Fig. 2a-c).

**Fig. 2.**
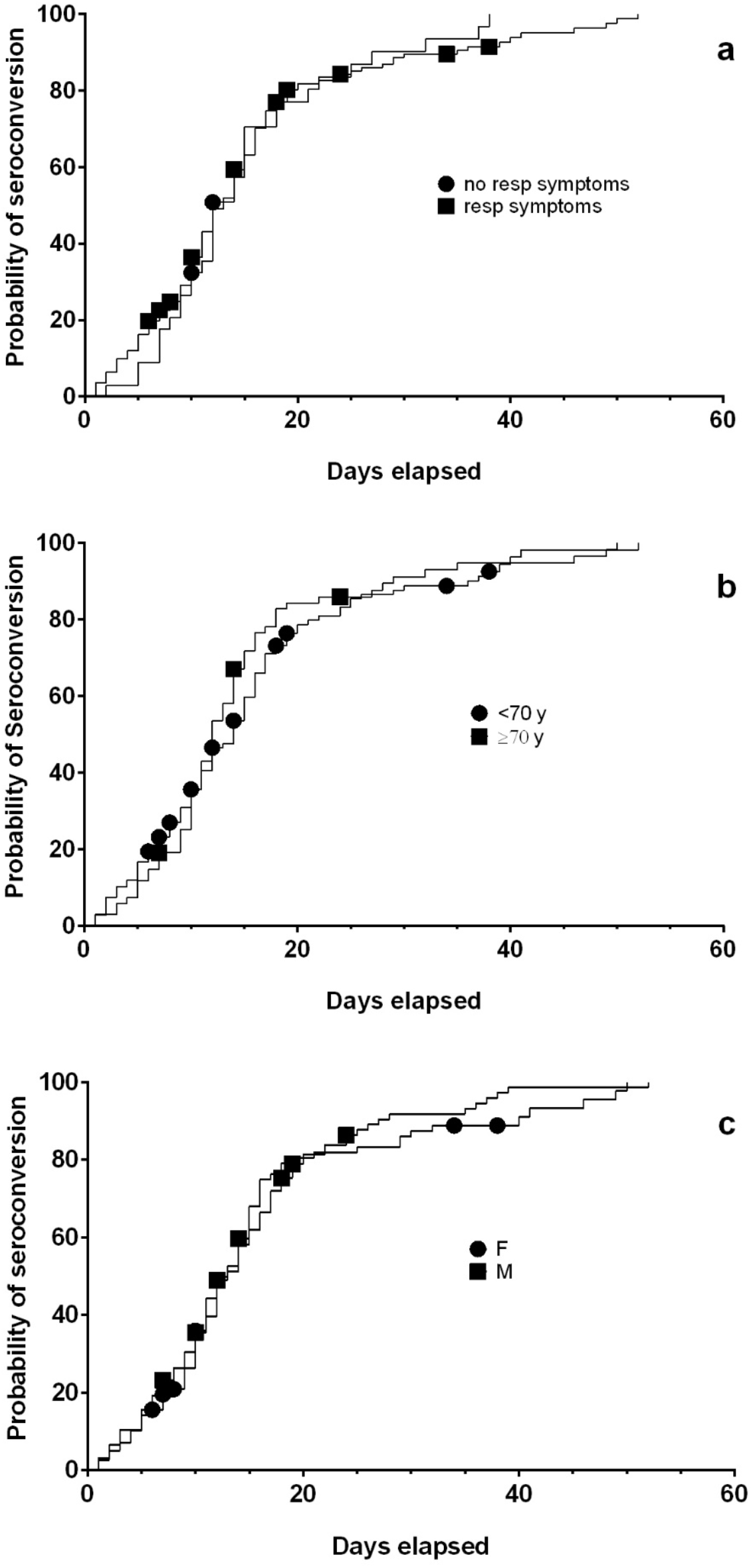
Cumulative frequency plots for seroconversion. Probability of seroconversion is described in days after an RT-PCR positive swab. **a**, Patients presenting with (filled squares) or without (filled circles) respiratory symptoms are shown, with the last available seronegative sample for a patient indicated by symbols and subsequently right censored. **b**, Patients aged ≥ 70 y (filled squares) or < 70 y (filled circles) are shown, with the last available seronegative sample for a patient indicated by symbols and subsequently right censored. **c**, Patients male (filled squares) or female (filled circles) are shown, with the last available seronegative sample for a patient indicated by symbols and subsequently right censored.

NOD IgG levels were not influenced by sex or presence or absence of respiratory symptoms (Supplementary Fig. 2a,b). Non-white ethnicity was associated with higher NOD levels than being white (mean values 1.06 vs 0.85 respectively;p = 0.035, F = 1.61, df = 119; unpaired Student’s *t*-test; Supplementary Fig. 2c).

Seroconverters (Fig. 1a) were older than non-seroconverters (median age 65.5 vs 41 years; p = 0.0033; Mann Whitney test) and were more likely to have one or more co-morbidities (124/130 vs 38/47; p = 0.0045; Fisher’s exact test). Hypertension was associated with a higher probability of seroconversion (74/75 hypertensives seroconverted vs 88/102 non-hypertensives;p = 0.0025; Fisher’s exact test). Body mass index (BMI) was higher among the group who seroconverted (25.7 vs 21.2; p = 0.034; Mann Whitney test).

C-reactive protein (CRP) is measured routinely in patients with COVID-19 unlike other markers of inflammation, and rising CRP values are poor prognostic indicators that are associated with cytokine release syndrome, if other causes are excluded^5,6^. CRP values on admission were significantly higher in patients with respiratory symptoms compared with those without (Fig. 3a). Patients with poor outcomes (death or admission to ITU) had higher CRP values than patients with good outcomes (Fig. 3b). Interestingly, patients who did not seroconvert had lower CRP values than seroconverters (Fig. 3c). The CRP differences by outcomes and seroconversion were more marked when the peak CRP values were analysed (Fig. 3m right hand panel). Other inflammatory markers, such as peak D-dimer, fibrinogen or ferritin were also higher in symptomatic compared with asymptomatic patients, but were available for fewer patients (Table 2).

**Table 2.** Laboratory values obtained at time of RT-PCR diagnosis and peak recorded values.

| **Test (normal range)** | **At time of RT-PCR**<br>**Median (IQR)** | **Peak value**<br>**Median (IQR)** |
| --- | --- | --- |
| CRP<br>(0 - 5 mg/L) | 86 (52.5 - 164)<br>N = 134 | 215.5 (103 - 334)<br>N = 176 |
| Nadir lymphocytes count<br>(1.1 - 4 x 10<sup>9</sup>/L) | 0.9 (0.6 - 1.4)<br>N = 134 | 0.6 (0.4 - 0.8)<br>N = 177 |
| Ferritin<br>(30 - 400 µg/L) | 1084 (630 - 1721)<br>N = 42 | 1335 (846 - 2758)<br>N = 89 |
| Fibrinogen<br>(1.6 - 4.8 g/L) | 5.5 (4.2 - 6.7)<br>N = 133 | 7.0 (5.5 - 9.15)<br>N = 166 |
| D-dimer<br>(21 - 300 ng/ml) | 704 (395 - 1078.75)<br>N = 64 | 1905 (498 - 4095)<br>N = 111 |
| Lactate dehydrogenase<br>(0 - 250 U/L) | 475 (280 - 597.5)<br>N = 47 | 490 (344.5 - 704)<br>N = 87 |

**Fig. 3.**
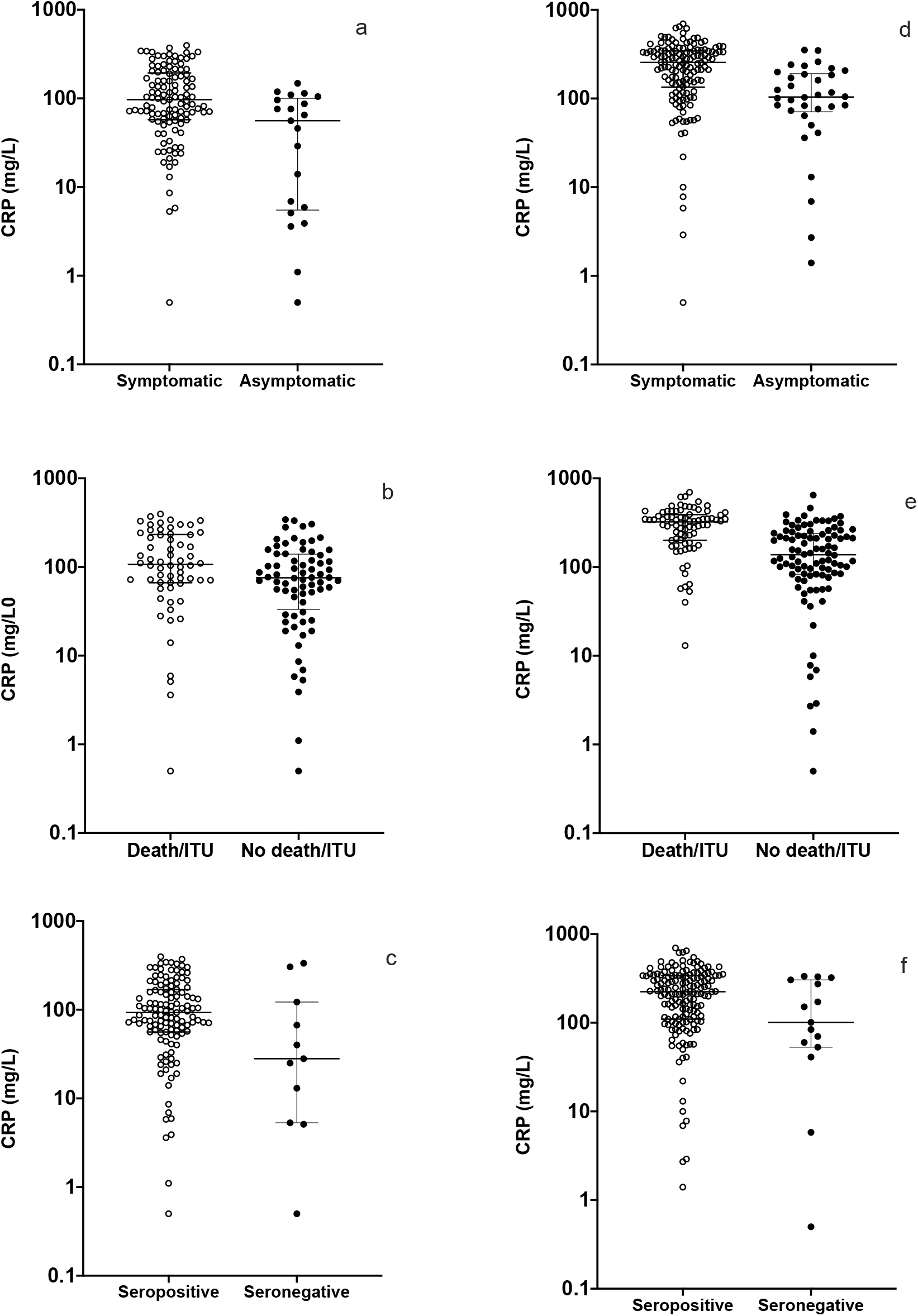
Relationships between CRP values, symptoms, outcomes, and NOD values. CRP levels (left hand panel; medians and interquartile ranges) at the time of the RT-PCR diagnosis are shown for **a**, symptomatic (open circles) and asymptomatic (filled circles) patients; **b**, for patients admitted to intensive care and/or who died (open circles) and those who were not (closed circles); **c**, those who seroconverted (open circles) and those who did not (closed circles). **d-f**, Peak CRP levels corresponding to **a-c** are shown (right hand panel; medians and interquartile ranges). Statistical significance calculated using Mann-Whitney test for CRP values (mg/L). **a**, 97 (n = 113) vs 56 (n = 21), p = 0.0008; **b**, 107 (n = 62) vs 75.5 (n = 72), p = 0.011; **c**, 93 (n = 123) vs 28 (n = 11), p = 0.04; **d**, median values respectively of 255 (n = 142) compared with 104 (n = 34) respectively, p < 0.0001; **e**, 322 (n = 80) vs 137.5 (n = 96), p < 0.0001; **f**, 224 (n = 161) vs 101 (n = 15), p = 0.025.

## Discussion

Serological tests for SARS-CoV-2 infections can enhance diagnostic capability for case management especially when the sensitivity of RT-PCR to diagnose COVID-19 falls later in the course of an infection^4,7,8^. We show dynamics of serological responses in patients presenting to hospital after many days of symptoms, when viral loads may be decreasing. Serological testing can also inform infection control measures allowing a better awareness of the potential infection risk of patients and their immune status and it can facilitate sero-epidemiological studies aiding contact tracing. Our study highlights that a significant proportion of patients with COVID-19 require several weeks post-infection with SARS-CoV-2 to generate antibodies. Furthermore, a proportion (2 - 8.5%) of patients do not have detectable antibodies up to 60 days post infection. Hitherto, most experience on antibody dynamics has come from investigators involved in the early response in China, reflecting the temporal unfolding of the pandemic^4,7,9^. Here, we describe variables that influence IgG antibody dynamics in SARS-CoV-2 infections in diverse populations.

The performance metrics of the IgG ELISA used here (Fig. 1 and Supplementary Table 1) after assessments on some of the largest COVID-19 datasets are comparable to other validated assays. This first-generation ELISA appears to have clinical utility to confirm an infection in patients without virological information. We have applied this test to study an ethnically and clinically diverse population and included individuals with asymptomatic, severe and fatal SARS-CoV-2 infections. NOD values remained stable for weeks after infection in most individuals who seroconverted, and seroconversion was relatively rapid (Fig. 1). The probability of seroconversion was associated with symptomatic disease, increased age, and presence of co-morbidities such as hypertension and increased BMI. Higher NOD values were associated with ethnicity (being non-white), probability of admission to hospital, and higher peak values for markers of inflammation such as CRP. Higher antibody titres are associated with clinical severity^4^.

CRP is a sensitive marker of elevated pro-inflammatory cytokines like Interleukin-6 that are hypothesised to play a central role in the cytokine release syndrome that presages a high mortality^6^. Interventions that target the proinflammatory cascade such as the anti-interleukin-6 receptor antibody, tocilizumab, may reverse disease progression and reduce mortality^10,11^ and are being studied urgently in randomised clinical trials (20 trials listed currently). A small proportion of individuals did not seroconvert even by 20 days after a positive RT-PCR test for SARS-CoV2. It may be that these patients never seroconvert or they may have immune responses confined to other antigens or mediated through T cells. Another likely explanation is that relatively mild infections may have been restricted to the mucosal cells of the respiratory tract, where antibody responses are dominated by the secretory immune system and little, if any, IgG antibody production in the systemic immune compartment.

The association of higher NOD ELISA values with elevated CRP could be interpreted in several ways. It is possible that antibody responses are working in concert with the cytokine response syndrome and are associated with more severe disease and risk of death through this mechanism. Alternatively, elevated CRP values may indicate a more marked innate response to infection in those at risk of severe disease and death. This heightened innate response may itself be associated with a higher viral load (perhaps through enhanced replication mechanisms) and interactions with genes that influence innate inflammatory pathways. A higher viral load would be predicted to give rise to higher NOD ELISA values for antibodies in the acquired immune response pathways. The potential therapeutic benefits of passive antibody transfer studies (in small trials^12^) suggest that the latter hypothesis is more likely^13^. Higher antibody responses are also seen with higher doses of non-replicating Ad5 vectored SARS-CoV-2 vaccine^14^.

Limitations of RT-PCR testing include difficulties with sampling, with different sample types and techniques yielding variable results, and diminishing diagnostic yield after peak viral loads that usually occur at the time of symptoms^9,15^. There may also be biological false positives associated with persistence of viral nucleic acid but no longer infective virus weeks after infection, or in sample handling. Serological testing, and the ability to detect viral antigens, may increase diagnostic accuracy for COVID-19 and our findings support early studies suggesting that these diagnostic modalities should be combined, particularly when RT-PCR results are negative but patients’ symptoms resemble those of COVID-19^7^. Late presentation to secondary care is common and may be part of containment strategies that encourage isolation.

One limitation of our study is that it is based mainly on hospitalised patients of whom 1 in 5 did not present with symptoms of COVID-19. Further studies are needed to document antibody dynamics of patients with less severe infections, such as in healthcare workers in eastern France^16^ and those who may have been missed because they already had low viral loads at the time of consultation. However, our findings will complement the large cross-sectional and longitudinal serological surveys being implemented as high quality tests become more widely available. ELISA NOD values were within a limited dynamic range (dilution studies could not be carried out because sample volumes or resources were limiting) but, nevertheless, were found to be related to clinically relevant features of COVID-19. Prospective studies are assessing the relationships between viral loads (assessed by RT-PCR) and serological responses in patients. Regular and longer-term serological assays will also be needed to monitor the duration and contribution of humoral responses to protection from SARS-CoV-2.

## Methods

### Ethics

Development of the SARS-CoV-2 IgG ELISA assay is published elsewhere^17^. Anonymized excess diagnostic material (EDM) from patients with SARS-CoV-2 infection confirmed by RT-PCR were used to evaluate antibody dynamics in these individuals. The study is sponsored by St George’s Hospital NHS Trust Foundation and has Institutional Review Board ethical approval (DARTS study - IRAS project ID: 282104; REC reference: 20/SC/0171). The trial is registered at clinicaltrials.gov NCT04351646.

### Reference RT-PCR

RT-PCR confirmation of SARS-CoV-2 infection was on samples from nose/throat swabs (in Sigma Virocult®, Corsham, UK) and Roche RNA extraction kits (Magnapure, West Sussex, UK) followed by altona Diagnostics RealStar® SARS-CoV-2 RT-PCR Kit (S and E genes, Hamburg, Germany) or Roche cobas® SARS-CoV-2 Test (E and ORF targets).

### Clinical samples

Serum samples from SARS-CoV-2 RT-PCR-positive subjects taken for clinical management at South West London Pathology (SWLP) provided EDM that was anonymised before analysis. Patients were sampled longitudinally to assess antibody dynamics and a minimum of 30 samples per day was included. If samples became unavailable from one patient, a new patient was added to the cohort. EDM was available from 177 patients between March 29^th^ and May 22^nd^ 2020 (inclusive). The study population was 9.9% (177/1785) of all SARS-CoV-2 patients at SWLP during the study period. St George’s Hospital is a tertiary teaching hospital and regional laboratory for microbial diagnostics.

### Participants and clinical data

Data were obtained from patients’ electronic medical records. Available outcomes were coded as hospital admission, Intensive Therapy Unit (ITU) stay, and death or discharge from hospital by May 22^nd^ 2020. Length of hospital stay was recorded for those who were discharged or died. Peak values of inflammatory markers (e.g. C-Reactive Protein, CRP) were the highest values recorded between 5 days prior to the first SARS-CoV-2 confirmation to the end of the study. Blood values at the time of the first RT-PCR positive result were obtained within 3 days of result notification to the clinical services (either way).

### Enzyme-linked immunosorbent assay (ELISA) for human IgG

The COVID-19 IgG ELISA assay developed by Mologic (Bedford, UK) and manufactured by Omega (Omega Diagnostics, Cambridge UK) was used according to the manufacturer’s instructions (Supplementary Methods). Between plate coefficients of variation were: lower cutoff 21% and positive control 16.5% (n = 16). Higher ambient temperatures gave higher readings. Performance characteristics of this assay are in Supplementary Table 1 (which was partly published^17^ previously and is provided for ease of access).

### Statistical analyses

Raw ELISA data were cross checked by 2 researchers and normalized (Supplementary Methods 1.2 and Figure 1.1) to allow comparison of data between ELISA plates. Manual handling errors were resolved as described in Supplementary Methods 1.3. Two-tailed parametric and non-parametric tests were applied as appropriate, with data analysis and display in PRISM (v8).

## Data Availability

Supplementary data for methods and results referred to in the manuscript is submitted together with the manuscript.

## Acknowledgements

We are greatly indebted to the National Association of Blood Bikes (NABB) for their unwavering assistance in transporting samples, prototypes, and validated devices between Liverpool, Bedford, and London. Without the NABB, we could not have delivered two CE marked products in 8 weeks from project launch. We also thank the tireless laboratory staff of SWLP and informatics team particularly James Lawrence.

## Data availability

The data that support the findings of this study are available from the corresponding authors upon reasonable request.

## Authors’ contributions

Conceptualisation: HMS, ERA, LEC, JRAF, SK, TP. Methodology: HMS, DEK, YA, MC, DF, SK,TP. Investigation: HMS, DEK, DJC, RLB, MC, ACA, BMOD, AD, NME, AJF, GG, LH, QH, MJ, GAK, KK, ZL, JMK, IM, CMM, GNC, SIO, CS, JS, CW, JW, KW. Supervision: HMS, DJC, ERA, LEC, MD, PD, DCM, AAS, JRAF, SK, TP. Formal Analysis: HMS, DEK, DJC, JMK, JRAF, SK, TP. Writing – Original Draft: HMS, DEK, DJC, SK, TP. Writing – Review & Editing: All. Funding Acquisition: HMS, ERA, LEC, AAS, JRAF, SK.

## Supplement

### Supplementary Methods

#### 1.1 ELISA

Serum samples were diluted one in 200 in dilution solution (10 mM Tris buffered saline, pH 7.2 with antimicrobial agents) and 100 µl loaded per well, in duplicate (although an initial plate was loaded in triplicate), onto a 96 well microtitre plate coated with SARS-CoV-2 antigens, spike, S, and nucleo-proteins, NP (an early generation assay used NP, S1 and S2 antigens and the finalized version used only NP and S2 antigens, with S1 being removed). Six wells contained controls: negative (diluent solution), negative cut-off (purified human IgG in 10 mM Tris buffered saline) and positive (purified human IgG in 10 mM Tris buffered saline), in duplicates. The plate was incubated at room temperature for 30 min prior to 3 washes with wash buffer (10mM Tris-buffered saline with detergent, pH 7.2). Following the wash step, 100 µl of conjugate solution (anti-Human IgG conjugated to horse radish peroxidase with protein stabilisation and antimicrobial agents) was added to every well and the plate incubated for 30 min at room temperature prior to 4 washes with wash buffer. Indicator/substrate solution (3,3′,5,5′-tetramethylbenzidine, TMB, with H_2_O_2_; 100 µl) was added to every well and the plate incubated at room temperature for 10 min prior to addition of 100 µl stop solution (0.25 M H_2_SO_4_) per well. The plate was read with a spectrometer at 450 nm within 10 min of stop solution addition. Wells with an optical density of 10% greater than the negative cut-off value were regarded as positive for antibodies to SARS-CoV-2 antigen.

#### 1.2 Normalisation

For each plate the mean cut off OD value plus 10% (lower cutoff) was subtracted from each mean patient sample OD value (thus samples with negative values were considered negative and samples with positive values were considered positive for SARS-CoV-2 antibodies). To normalize, the resulting value was divided by the mean positive control OD value. Any assayed duplicates (serum samples taken from the same patient on the same day) were removed from further analysis.

#### 1.3 Resolving inconsistencies

Inconsistent data points (n = 3) were identified (*e*.*g*. a data point that suggested a seropositive patient subsequently lost their antibody response). In these cases, samples were re-run including sequential samples from the same patient from either side of the timepoint, alkaline phosphatase results from that sample were checked and sample aliquots were resourced from stocks in SWLP laboratories.

### Supplementary Results

#### 1.1 Normalised ELISA values

Normalised OD values for ELISA results are presented in Supplementary Figure 1

**Supplementary Fig. 1.**
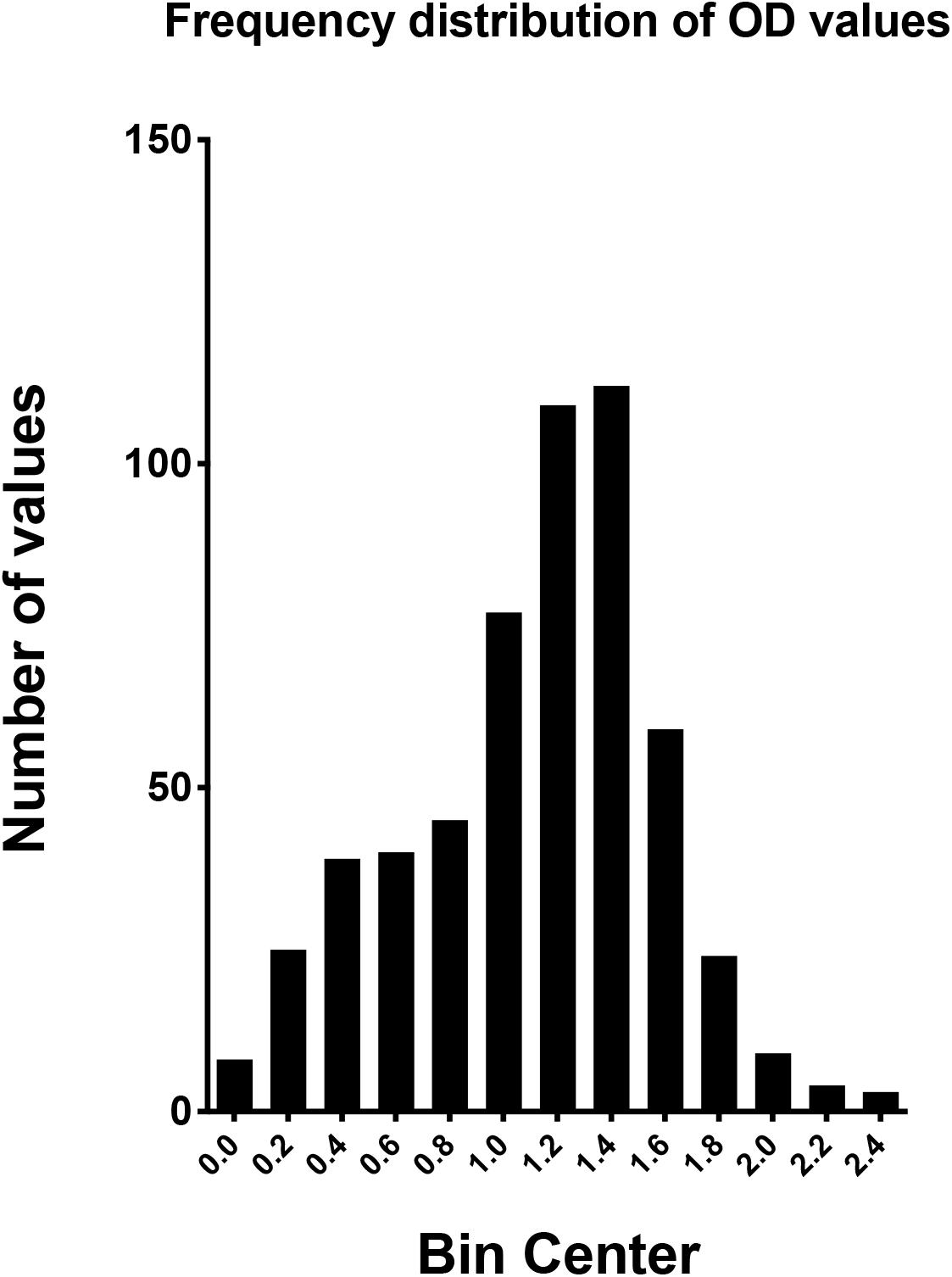
Raw ELISA values were normalized as described in Supplementary methods 1.2. This distribution passed (p > 0.05) all normality tests offered in PRISM.

**Supplementary Fig. 2.**
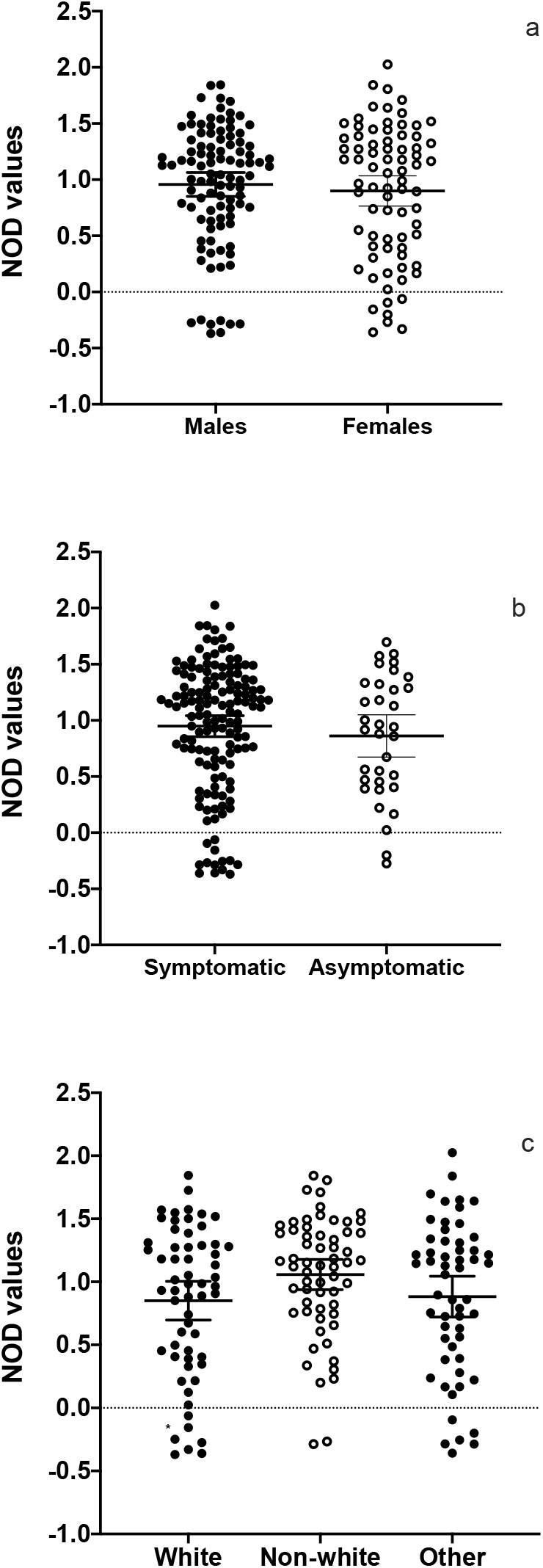
NOD values compared by sex, symptoms and ethnicity. **a**, Comparison of NOD values by sex (mean ± 95% CI; not significantly different);**b**, Comparison of NOD values by symptoms (mean ± 95% CI;not significantly different); **c** Comparison of NOD values by ethnicity (mean ± 95% CI; p = 0.035, F = 1.61, df = 119; unpaired Student’s *t*-test between white and non-white). The third group is not known/other.

**Supplementary Table 1.**
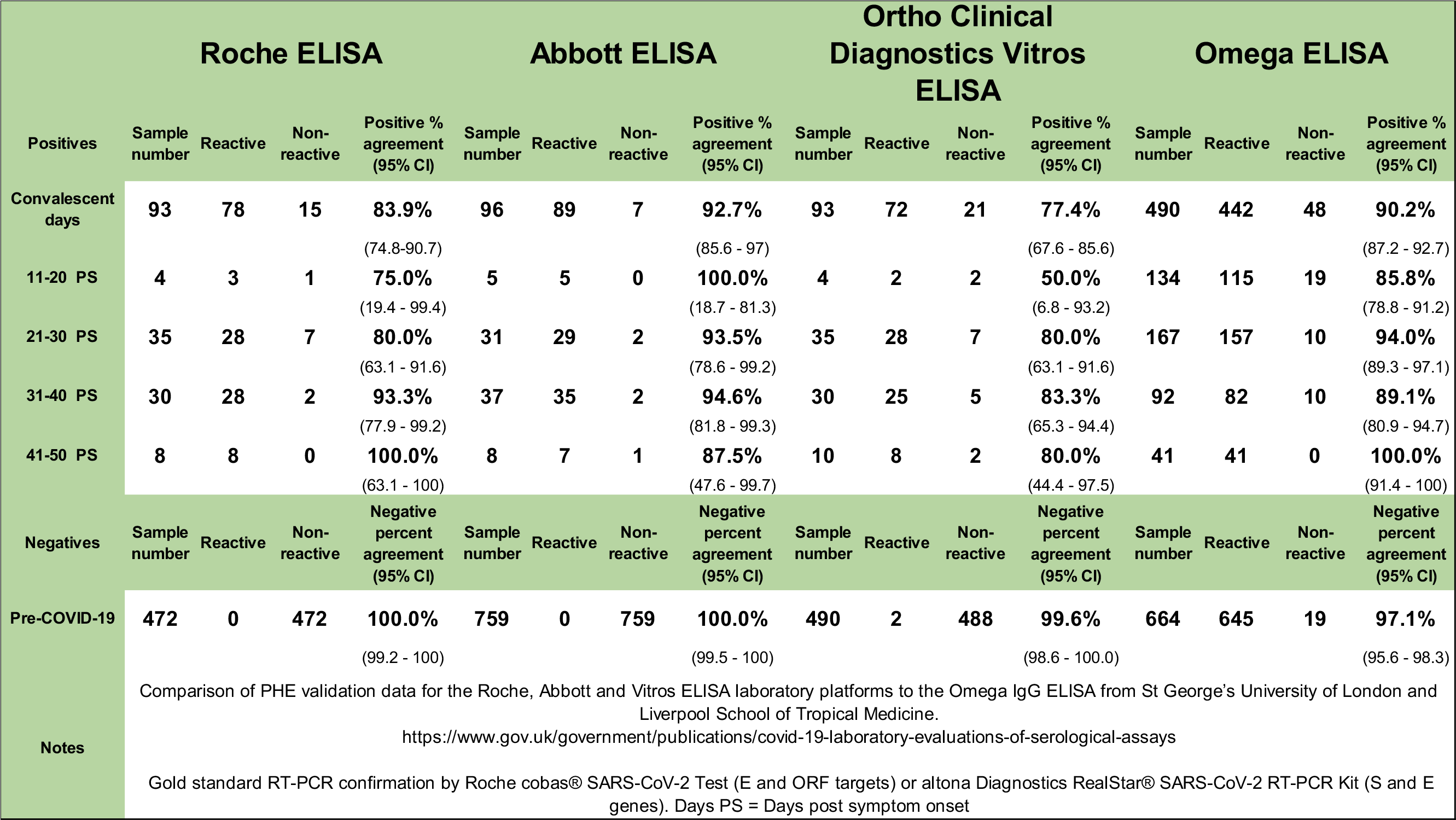
Comparison of commercially-available, independently validated ELISA platforms for SARS-CoV-2 Immunoglobulin G.

**Supplementary Table 2.** Expanded characteristics of study patients.

|  | **Total N = 177 (%)** |
| --- | --- |
| **Ethnicity** |  |
| **White** | 60 (33.9) |
| White British | 38 (21.5) |
| White Irish | 1 (0.6) |
| White - Any Other White Background | 21 (11.9) |
| **Non-white** | 61 (34.5) |
| Black or Black British - African | 13 (7.3) |
| Black or Black British - Caribbean | 9 (5.1) |
| Black - Any Other Black Background | 5 (2.8) |
| Asian or Asian British - Bangladeshi | 1 (0.6) |
| Asian or Asian British - Indian | 6 (3.4) |
| Asian or Asian British - Pakistani | 2 (1.1) |
| Asian - any other Asian background | 22 (12.4) |
| Mixed - Any Other Mixed Background | 3 (1.7) |
| **Other/not known** | 56 (31.6) |
| Other - any other ethnic group | 33 (18.6) |
| Other - not stated | 23 (13) |
| **Comorbidities** |  |
| **None** | 47 (26.6) |
| Diabetes mellitus | 52 (29.4) |
| Hypertension | 75 (42.4) |
| Obesity (BMI >30)* | 44/164 (26.8) |
| Chronic kidney disease | 27 (15.3) |
| CKD | 15 (8.5) |
| Haemodialysis | 10 (5.7) |
| Renal transplant | 2 (1.1) |
| Cancer | 18 (10.2) |
| Asthma | 21 (11.9) |
| COPD | 6 (3.4) |

Ethnicity data are routinely collected by the National Health Service and categorised based on the 2001 UK Census classification. \*Height data unavailable for 13 patients.

## References

1. Yang, X., et al. Clinical course and outcomes of critically ill patients with SARS-CoV-2 pneumonia in Wuhan, China: a single-centered, retrospective, observational study. Lancet Respir Med 8, 475-481 (2020).

2. Chan, J.F., et al. A familial cluster of pneumonia associated with the 2019 novel coronavirus indicating person-to-person transmission: a study of a family cluster. Lancet 395, 514-523 (2020).

3. https://ourworldindata.org/coronavirus

4. Zhao, J., et al. Antibody responses to SARS-CoV-2 in patients of novel coronavirus disease 2019. Clin Infect Dis (2020).

5. Ruan, Q., Yang, K., Wang, W., Jiang, L. & Song, J. Correction to: Clinical predictors of mortality due to COVID-19 based on an analysis of data of 150 patients from Wuhan, China. Intensive Care Med (2020).

6. Woo, Y.L., et al. A genetic predisposition for Cytokine Storm in life-threatening COVID-19 infection. OSF Preprints, mxsvw (2020).

7. Guo, L., et al. Profiling Early Humoral Response to Diagnose Novel Coronavirus Disease (COVID-19). Clin Infect Dis (2020).

8. Kucirka, L.M., Lauer, S.A., Laeyendecker, O., Boon, D. & Lessler, J. Variation in False-Negative Rate of Reverse Transcriptase Polymerase Chain Reaction-Based SARS-CoV-2 Tests by Time Since Exposure. Ann Intern Med (2020).

9. Long, Q.X., et al. Antibody responses to SARS-CoV-2 in patients with COVID-19. Nat Med (2020).

10. Baker, E.H., et al. Response to Tocilizumab Treatment in Severe COVID-19. OSF Preprints, d2nh8 (2020).

11. Xu, X., et al. Effective treatment of severe COVID-19 patients with tocilizumab. Proc Natl Acad Sci U S A 117, 10970-10975 (2020).

12. Salazar, E., et al. Treatment of COVID-19 Patients with Convalescent Plasma. Am J Pathol (2020).

13. Jiang, S., Hillyer, C. & Du, L. Neutralizing Antibodies against SARS-CoV-2 and Other Human Coronaviruses: (Trends in Immunology 41, 355-359m 2020). Trends Immunol 41, 545 (2020).

14. Zhu, F.C., et al. Safety, tolerability, and immunogenicity of a recombinant adenovirus type-5 vectored COVID-19 vaccine: a dose-escalation, open-label, non-randomised, first-in-human trial. Lancet (2020).

15. Xie, C., et al. Comparison of different samples for 2019 novel coronavirus detection by nucleic acid amplification tests. Int J Infect Dis 93, 264-267 (2020).

16. Fafi-Kremer, S., et al. Serologic responses to SARS-CoV-2 infection among hospital staff with mild disease in eastern France. medRxiv, 2020.2005.2019.20101832 (2020).

17. Adams, E.R., et al. Rapid development of COVID-19 rapid diagnostics for low resource settings: accelerating delivery through transparency, responsiveness, and open collaboration. medRxiv, 2020.2004.2029.20082099 (2020).

